# Physical activity and fiber intake beneficial for muscle mass and strength preservation during aging: A Comprehensive Cross-Sectional Study in the UK Biobank cohort

**DOI:** 10.1101/2024.01.22.23300657

**Authors:** Jordi Morwani-Mangnani, Mar Rodriguez-Girondo, Cecile Singh-Povel, Sjors Verlaan, Marian Beekman, P. Eline Slagboom

**Affiliations:** Section of Molecular Epidemiology, Department of Biomedical Data Sciences, Leiden University Medical Center, Leiden, Netherlands; FrieslandCampina, Amersfoort, The Netherlands

**Keywords:** healthy aging, biology of aging, lifestyle, nutrition, fiber intake, muscle health, UK Biobank

## Abstract

**Background:** Aging triggers intricate physiological changes, particularly in muscle mass and strength, affecting overall health and independence. Despite existing research, the broader significance of how muscle health is affected by the intricate interplay of lifestyle factors simultaneously during aging needs more exploration. This study aims to examine how nutrition, exercise, and sleep independently and differentially impact on muscle mass and strength in middle-aged men and women, facilitating future personalized recommendations for preserving muscle health.

**Methods:** The cross-sectional analysis of the UK Biobank involved 45,984 individuals aged 40-70. Multiple linear regression explored determinants of whole-body fat-free mass and handgrip strength, considering traditional, socio-demographics, medication use and smoking as covariates, with gender and age (younger and older than 55 years) stratifications.

**Results:** Higher physical activity and fiber intake beneficially affect both muscle mass and strength, especially above 55 years. Other lifestyle factors influence the two muscle parameters differently. Key determinants influencing muscle strength included higher protein intake, lower water intake, higher alcohol intake, and extended sleep duration whereas mainly higher water intake contributes to higher muscle mass.

**Conclusions:** Physical activity and fiber intake associate with increased muscle strength and mass which may connect gut and muscle health. Given the observed complexity of muscle health in the age and gender strata, further longitudinal research is needed to provide personalized lifestyle recommendations.

## INTRODUCTION

Aging consists of a complex variety of physiological changes in all tissues. Specifically, shifts in muscle mass and strength eventually contributing to sarcopenia have a significant impact on health and independent functioning at older ages.^1,2^ Changes in muscle mass and strength, however, do not always decline in parallel as we age. Variations in muscle strength often appear before significant reductions in muscle mass, which highlights the need to study the unique biological factors controlling these aspects separately. The muscle changes determine the individual health span and are largely influenced by sociodemographic and lifestyle factors.^3,4^

The relationship between muscle health and traditional, clinical, lifestyle, and socio-demographic factors as previously explored is displayed in TABLE S1. Variables such as older age, female gender, lower BMI, lower waist circumference, high medication use, smoking, alcohol consumption, lower socioeconomic status, and Asian ethnicity are associated with lower muscle mass and strength.^4,13–18^ In addition, recent research has emphasized the impact also of other lifestyle factors like macronutrient intake and physical activity. Factors such as higher protein intake, water intake, dietary fiber, physical activity levels, and at least 7 hours of sleep have all been investigated for their separate positive effects on muscle health.^3–8^ Sufficient water intake is important to safeguard muscle cell volume and function while alcohol consumption is associated with myopathies characterized by lower muscle protein content.^4,9,10^

Until now, most studies have examined all these effects individually, leading to a disjointed comprehension of the effect of lifestyle on muscle health. Understanding the intricate interactions affecting muscle health is crucial in shaping robust, age- and or sex-specific strategies to enforce muscle health in a personalized fashion.

This study aims to understand how nutrition (including protein, fiber, water, alcohol intake), exercise, and sleep have independent and differential effects on muscle mass and strength in younger (≤55 years) and older (>55) men and women of the UK-Biobank study. This study allows us to see how these factors interact with age- and gender-related changes, contributing to a more comprehensive understanding of muscle health throughout the lifespan.

## METHODS

### Study design

The present cross-sectional study draws from the UK Biobank, a substantial cohort study encompassing a diverse array of data sources.^11^ From the initial 502,628 participants enrolled at baseline, a cohort of 45,984 individuals (comprising 20,510 men and 25,474 women) met the inclusion criteria for this investigation (TABLE 1). Specifically, these participants were aged 40-70 years and possessed complete datasets for the outcomes under study, exposures pertaining to lifestyle factors, and the array of covariates of interest, including traditional covariates, socio-demographic variables, medication utilization, and smoking status (TABLE S1). Ethical clearance for this research was acquired under the broad ethical approval secured by UK Biobank investigators from the National Health Service National Research Ethics Service. The present analyses were conducted in accordance with application number 78275.

**TABLE 1.** Baseline characteristics of the study populations for men and women younger and older than 55 years. Sleepers were categorized according to sleep duration: short sleepers <6.9hours; normal sleepers 7-9 hours; long sleepers >9hour. *This variable is for exploratory purposes only and will not be included in the analyses.

|  | Men |  | Women |  | All |
| --- | --- | --- | --- | --- | --- |
|  | ≤55<br>(N=8903) | >55<br>(N=12 123) | ≤55<br>(N=11607) | >55<br>(N=13351) | (N=45984) |
| **Outcomes of Interest** |  |  |  |  |  |
| Whole Body Fat Free Mass (WBFFM, kg), mean (SD) | 65.5 (7.91) | 63.0 (7.35) | 45.4 (5.10) | 43.9 (4.71) | 53.5 (11.6) |
| Handgrip strength (HGS, kg), mean (SD) | 41.1 (8.69) | 37.0 (7.91) | 25.1 (5.88) | 21.7 (5.56) | 30.4 (10.6) |
| **Lifestyle Factors** |  |  |  |  |  |
| Physical activity (hours/week), mean (SD) | 46.8 (49.1) | 46.1 (45.8) | 42.8 (41.0) | 45.2 (41.5) | 45.1 (44.1) |
| Fibre Adjusted Intake (g/day), mean (SD) | 15.3 (6.99) | 16.7 (7.00) | 16.8 (6.72) | 18.1 (6.67) | 16.9 (6.90) |
| Protein Intake (g/kgBW/day), mean (SD) * | 0.956<br>(0.304) | 0.977 (0.282) | 1.190 (0.350) | 1.210 (0.324) | 1.095 (0.337) |
| Protein Adjusted Intake (g/day), mean (SD) | 81.5 (23.5) | 81.8 (20.9) | 82.1 (19.8) | 83.2 (18.2) | 82.2 (20.4) |
| Water Intake(cups/day), mean (SD) | 3.13 (2.52) | 2.38 (1.98) | 3.42 (2.43) | 2.99 (2.10) | 2.96 (2.28) |
| Alcohol Adjusted Intake (g/day), mean (SD) | 19.9 (29.6) | 21.5 (27.9) | 13.0 (18.9) | 12.7 (17.0) | 16.5 (23.8) |
| **Sleep** |  |  |  |  |  |
| Short, n (%) | 6331<br>(71.1%) | 9310 (76.8%) | 8892 (76.6%) | 9925 (74.3%) | 34458 (74.9%) |
| Normal, n (%) | 2503<br>(28.1%) | 2639 (21.8%) | 2551 (22.0%) | 3239 (24.3%) | 10932 (23.8%) |
| Long, n (%) | 69 (0.8%) | 174 (1.4%) | 164 (1.4%) | 187 (1.4%) | 594 (1.3%) |
| **Traditional covariate** |  |  |  |  |  |
| Age (years), mean (SD) | 47.9 (4.62) | 62.6 (3.69) | 48.1 (4.54) | 62.1 (3.69) | 55.9 (8.20) |
| BMI, mean (SD) | 27.6 (4.27) | 27.6 (3.99) | 26.4 (5.25) | 26.8 (4.82) | 27.1 (4.66) |
| Waist circumference (cm), mean (SD) | 95.1 (11.3) | 97.2 (11.0) | 82.3 (12.5) | 84.8 (12.2) | 89.4 (13.4) |
| **Socio-demographic covariates** |  |  |  |  |  |
| **Assessment Centre** |  |  |  |  |  |
| Bristol, n (%) | 1271<br>(14.3%) | 1643 (13.6%) | 1716 (14.8%) | 1803 (13.5%) | 6433 (14.0%) |
| Sheffield, n (%) | 1717<br>(19.3%) | 2626 (21.7%) | 2188 (18.9%) | 2756 (20.6%) | 9287 (20.2%) |
| Liverpool, n (%) | 877 (9.9%) | 1513 (12.5%) | 1102 (9.5%) | 1575 (11.8%) | 5067 (11.0%) |
| Middlesborough, n (%) | 518 (5.8%) | 677 (5.6%) | 638 (5.5%) | 691 (5.2%) | 2524 (5.5%) |
| Hounslow, n (%) | 1646<br>(18.5%) | 2163 (17.8%) | 2161 (18.6%) | 2431 (18.2%) | 8401 (18.3%) |
| Croydon, n (%) | 1675<br>(18.8%) | 2123 (17.5%) | 2331 (20.1%) | 2588 (19.4%) | 8717 (19.0%) |
| Birmingham, n (%) | 1199<br>(13.5%) | 1378 (11.4%) | 1471 (12.7%) | 1507 (11.3%) | 5555 (12.1%) |
| **Ethnicity** |  |  |  |  |  |
| White, n (%) | 8165<br>(91.7%) | 11740 (96.8%) | 10608 (91.4%) | 12922 (96.8%) | 43435 (94.5%) |
| Mixed, n (%) | 94 (1.1%) | 29 (0.2%) | 174 (1.5%) | 47 (0.4%) | 344 (0.7%) |
| Asian, n (%) | 387 (4.3%) | 259 (2.1%) | 422 (3.6%) | 219 (1.6%) | 1287 (2.8%) |
| Black, n (%) | 257 (2.9%) | 95 (0.8%) | 403 (3.5%) | 163 (1.2%) | 918 (2.0%) |
| Index of Multiple Deprivation | 17.6 (12.9) | 15.1 (11.4) | 17.1 (12.5) | 15.2 (11.4) | 16.1 (12.0) |
| **Medication use** |  |  |  |  |  |
| Number medications, mean (SD) | 1.31 (1.87) | 2.53 (2.59) | 1.70 (2.06) | 2.49 (2.56) | 2.07 (2.38) |
| **Smoking Status, n (%)** |  |  |  |  |  |
| Smoker | 1097<br>(12.3%) | 907 (7.5%) | 1054 (9.1%) | 699 (5.2%) | 3757 (8.2%) |

### Study procedures

Methods of collection and data transformations for each measurement can be found in TABLE S1. Whole body fat-free mass (referred to as muscle mass throughout this article) was measured through the Tanita BC 418MA. Handgrip strength was measured once in each hand with a Jamar J00105 hydraulic hand dynamometer. In the analysis, we used the mean of both measurements.

### Statistical methods

To evaluate which set of lifestyle factors are independently associated with muscle mass and strength, we employed multiple linear regression. We constructed two models: The first with whole-body fat-free mass as an outcome (TABLE S2) and the second with handgrip strength as the outcome (TABLE S3). Both models included the seven lifestyle factors of interest, involving nutrition, physical activity, and sleep factors alongside traditional covariates such as age, gender, BMI, and waist circumference and socio-demographic factors such as assessment center (representing geographical distribution), ethnicity, and Townsend index of multiple deprivation.^12–14^ Additional covariates included the number of medications used and current smoking status. Adjustment for dietary variables in relation to energy intake was achieved using the Willett procedure to rectify residual errors.^15^ Gender-based stratification was performed, recognizing biological disparities, and further stratification grouped participants by younger and older age groups (also referred to as ≤55 and >55, respectively). The analysis adopted a Bonferroni correction with a significance threshold of p < 0.0036 to account for the 14 explanatory variables in the two models of whole-body fat-free mass and handgrip strength. Statistical analyses were performed using R version 4.2.3 and RStudio version 2023.06.1+524.

## RESULTS

### Study population

The cohort of 45,984 participants was stratified by gender and age categories to explore the biological and behavioral differences between them: 8,234 younger men, 12,783 older men, 10,761 younger women, and 14,197 older women (TABLE 1).

The participants in this study exhibited diverse characteristics, highlighting variations across four distinct age and gender categories (TABLE 1). All categories were evenly distributed among assessment centers, comprised of mainly white ethnicity, and displayed similar patterns in protein and fiber intake. Additionally, a substantial proportion of participants were identified as short sleepers. A gender-based comparison revealed that men exhibited higher muscle mass, handgrip strength, BMI, waist circumference, alcohol intake, marginally greater engagement in physical exercise, and slightly lower absolute protein intake and total-energy-intake-adjusted protein intake compared to women. When examining age-related differences, the subset younger than 55 years demonstrated higher muscle mass, handgrip strength, lower reliance on medication, a higher prevalence of smokers, lower fiber intake, and increased water consumption.

To study the independent associations of the known covariates of muscle mass and strengths, we performed multiple linear regressions with whole-body fat free mass (TABLE S2) and handgrip strength (TABLE S3) as outcomes while adjusting for age, body composition, socio-demographic and economic factors, medication use and smoking status. Below the most important outcomes are summarized.

### More physical activity is associated with both higher muscle mass and strength

Physical activity measured in MET hours per week had significant relevance in muscle parameters across gender and age groups (FIGURE 1, TABLE S2, TABLE S3). In older men and women, more physical activity was independently associated with higher muscle mass (respectively B = 3.36×10^−3^, p-value = 1.66×10^−3^; B = 2.52×10^−3^, p-value = 3.57×10^−4^) and muscle strength (respectively B = 6.05×10^−3^, p-value = 7.99×10^−5^, B = 8.98×10^−3^, p-value = 2.95×10^−15^). These associations are also reflected in men and women younger than 55 years for muscle mass (respectively B = 9.02×10^−3^, p-value = 1.45×10^−13^; B = 4.35×10^−3^, p-value = 4.11×10^−8^) and muscle strength (respectively B = 1.36×10^−2^, p-value = 1.00×10^−13^; B = 1.08×10^−2^, p-value = 8.52×10^−17^).

**FIGURE 1.**
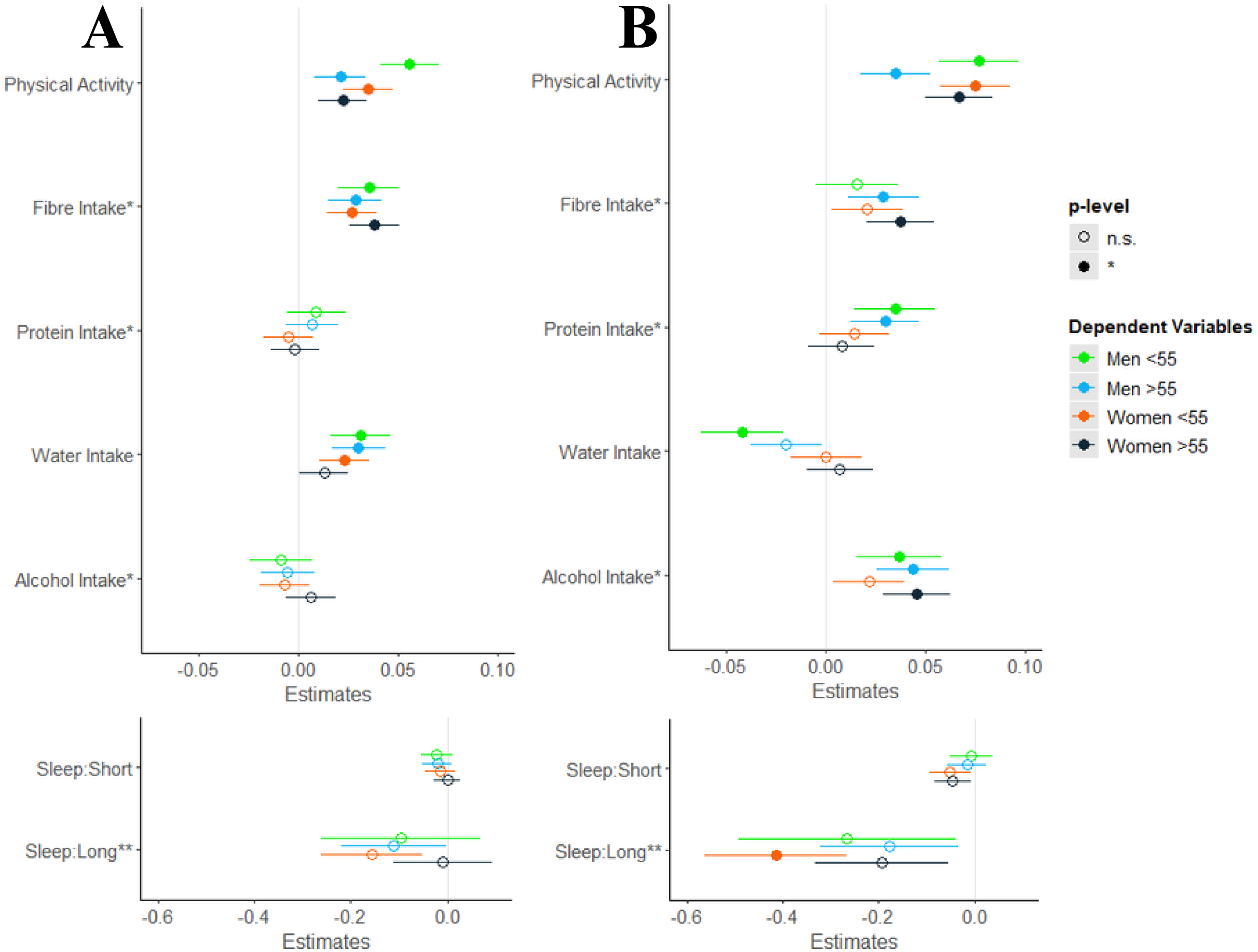
Lifestyle variables in the multivariate model presented as standardized beta-coefficients associated with. **(A)** muscle mass and **(B)** handgrip strength for men and women younger and older than 55 years. Adjusted for traditional covariates, socio-demographic covariates, medication use, and smoking status. The p-value at which to accept significance (filled in circle) was adjusted to account for multiple testing (p < 0.0036). Sleepers were categorized according to sleep duration: short sleepers <6.9hours; normal sleepers 7-9 hours; long sleepers >9hours. *Intakes adjusted for energy intake (see Methods). **Large confidence intervals reflect low statistical power for this variables.

### Higher fiber intake is associated with higher muscle mass and strength

Higher fiber intake was found to be associated with higher muscle mass and strength in various strata (FIGURE 1, TABLE S2, TABLE S3). In older men and women, higher fiber intake was associated with higher muscle mass (respectively B = 3.00×10^−2^, p-value = 2.76×10^−5^; B = 2.68×10^−2^, p-value = 1.78×10^−9^) and higher muscle strength (respectively B = 3.27×10^−2^, p-value = 1.40×10^−3^; B = 3.12 ×10^−2^, p-value = 1.34×10^−5^). In younger men and women, higher fiber intake was only significantly associated with higher muscle mass (respectively B = 4.01×10^−2^, p-value = 5.06×10^−6^; B = 2.04×10^−2^, p-value = 3.15×10^−5^).

### Higher protein intake is associated with higher muscle strength in men, but not women

Protein intake (adjusted for total energy intake) exhibited distinct associations with muscle parameters across gender and age groups (FIGURE 1, TABLE S2, TABLE S3). Specifically, protein intake (adjusted for total energy intake) showed no relation with muscle mass in men or women. However, it exhibited an association on muscle strength in both young and older men (respectively B = 1.29×10^−2^, p-value = 7.41×10^−4^; B = 1.13×10^−2^, p-value = 8.34×10^−4^). In contrast, protein intake (adjusted for total energy intake) showed no significant association with muscle strength nor mass, neither in young nor older women.

### Higher water intake is associated with higher muscle mass and not with muscle strength

Water intake had varying associations with muscle parameters across different subgroups (FIGURE 1, TABLE S2, TABLE S3). In men both younger and older than 55 years, higher water intake was associated with higher muscle mass (B = 9.89×10^−2^, p-value = 3.91×10^−5^; B = 1.13 ×10^−1^, p-value = 6.70 ×10^−6^, respectively). However, in men younger than 55 years, higher water intake associated with slightly lower muscle strength (B = -0.145, p-value = 6.19 ×10^−5^). Associations on muscle mass were observed in women younger than 55 years only (B = 4.86×10^−2^, p-value = 3.32 ×10^−4^).

### Higher alcohol intake is associated with higher muscle strength but not with muscle mass

Higher intake of alcohol did not associate with muscle mass but did with higher muscle strength in men both young and older than 55 years (B = 1.08×10^−2^, p-value = 5.67 ×10^−4^; B = 1.24×10^−2^, p-value = 1.63×10^−6^, respectively) and women older than 55 years (B = 1.49×10^−2^, p-value = 1.24×10^−7^) (FIGURE 1, TABLE S2, TABLE S3).

### Sleeping longer than 9 hours per night is associated with lower muscle strength in women younger than 55 years, but not in men

Sleep duration was found to have a unique impact on muscle parameters (FIGURE 1, TABLE S2, TABLE S3). While sleep duration showed no significant associations with muscle parameters in men, it was observed that sleeping longer than 9 hours was associated with lower muscle strength in women younger than 55 years(B = -2.439, p-value = 4.03 x 10^−8^).

## DISCUSSION

This study examined the link between lifestyle factors (diet, physical activity, and sleep) and muscle mass and strength in individuals aged 40-70 years. Multiple linear regression models revealed physical activity and fiber intake associated with both parameters. More factors independently influenced muscle strength than mass. Increased water intake contributed to increased muscle mass. Higher handgrip strength was influenced by higher protein intake, lower water intake, higher alcohol intake, and extended sleep duration. The study highlighted age and gender differences in the impact of lifestyle on muscle health in middle-aged to older adults.

The stratification based on age is a pivotal element of this study. It significantly enhances our comprehension of muscle health dynamics.^16^ The most remarkable observation is the attenuated effect of physical activity on muscle strength in men above 55 years, as compared to the other groups including older women. Muscle mass is related to physical activity similarly in both sexes over 55 years. It is essential to consider that gender- and age-related changes both play substantial roles in the development of sarcopenia, a condition characterized by loss of muscle mass and strength. With age, detrimental shifts occur in dietary practices and physical activity habits.^17,18^ It is important to consider the impact of behavioral differences on exercise in men and women. Men tend to engage more in strength training than women, while also adopting riskier behaviors fueled by intrinsic competitiveness. This may explain why men tend to focus on upper-body training for muscularity, while women focus on lower-body training and overall body mass management.^19,20^

Our results show that older men had the lowest muscle mass and strength out of the four groups. This observation aligns with existing research that posits a higher prevalence of sarcopenia in men compared to women, potentially elucidating the observed disparity in muscle parameters within our study.^28–30^ Beyond behavioral differences mentioned above, gender differences in muscle physiology exist. Young men have a larger volume distribution of type II muscle fibers, the main fiber type involved in strength training. Contrarily, women have a larger volume distribution of type I muscles, which are related to endurance training.^21,22^ Physical activity had the strongest association with muscle mass in men younger than 55 years as compared to the other groups. Indeed, physical activity has long been recognized for its positive impact on muscle health, with the present study reaffirming this well-established concept in both age and gender categories for muscle mass and strength. Regular exercise first stimulates the neuromuscular system, leading to increased motor unit recruitment and improved muscle contraction efficiency.^24^ Thereafter it elicits muscle hypertrophy, leading to an increase in muscle size, and promotes muscle protein synthesis, a fundamental process for muscle growth.^23^ However, muscle mass and strength decline with age, as confirmed in our data. This is especially observed in men, clearly not prevented by their physical activity. Our results therefore underscore that older individuals might need tailored exercise and dietary regimens to counteract these age-related changes and maintain or enhance their muscle mass and strength.^26,27^

The positive association of higher fiber intake with muscle mass across genders and age groups may be explained by the gut-muscle axis hypothesis, which posits that dietary fiber intake enhances gut microbiota diversity.^31–33^ A diverse gut microbiota promotes the production of short-chain fatty acids, which are known to influence muscle metabolism.^34^ Short-chain fatty acids regulate muscle protein synthesis and degradation, which in turn affect muscle mass.^35,36^ Further adding to the complexity of this relationship, our findings indicated age-based variations in the relationship between fiber intake and muscle strength highlighting the need for further research in this area.^37,38^ The literature corroborates our findings for both muscle mass and strength, where a higher fiber intake is correlated with higher lean mass and muscle strength.^39^

Protein intake, a widely recognized factor in muscle health, surprisingly did not show a significant association with muscle mass in our study. This could be attributed to our adjustment for total caloric intake (FIGURE S1), suggesting that muscle mass development does not solely rely on protein intake but also requires a balanced diet with appropriate caloric intake.^40,41^ Recent studies suggest that total-energy-unadjusted protein intake patterns across the day, and not only total protein intake, are essential for optimal muscle health.^42,43^ Furthermore, we found total-energy-intake-adjusted protein intake to be associated with increased muscle strength in men, but not in women. This gender-based variability in response to protein intake might be due to, men having more lean mass than women and inherently more strength, requiring a higher absolute protein intake to preserve their muscle health.^44^ We observed a similar total-energy-adjusted protein intake among men and women. However, the absence of effect of protein intake on muscle mass and strength in women may be due to the visibly higher SD compared to men.

Higher water intake was found to associate with higher muscle mass, particularly in men in both age categories and women younger than 55 years. Hydration status can also influence muscle cell volume, with dehydration potentially leading to muscle atrophy due to the loss of intracellular water.^7^ Conversely, our study showed that higher water intake is associated with lower muscle strength in men younger than 55 years. Overhydration can disrupt electrolyte balance, potentially leading to muscle weakness due to impaired muscle contraction.^46^ Previous studies support the importance of hydration for both muscle mass and strength, especially in older adults.^47^ This age-based difference underscores the importance of individualized hydration strategies for optimal muscle strength.

Women older than 55 years had the lowest alcohol intake and the highest protective effect on muscle strength. The observed protective effect of alcohol specifically on muscle strength in men of both age categories and women older than 55 years is intriguing and can be understood through several interconnected mechanisms. Firstly, limited alcohol consumption has been shown to provide neuroprotection through its antioxidative and anti-inflammatory properties.^48^ Alcohol, particularly at modest doses, exhibits antioxidant capabilities, which can counteract the damaging effects of oxidative stress and inflammation.^49^ This, in turn, can safeguard neuronal function, contributing to enhanced muscle control and strength. Gender differences in behaviors in alcohol consumption may underpin the absence of significance in younger women.^50^ Overall, the present results are consistent with previous explorations regarding alcohol intake and muscle health within the same cohort.^51^

Lastly, we show that sleeping longer than 9 ours is associated with lower muscle strength. This unexpected finding suggests that there might be underlying physiological mechanisms, such as hormonal fluctuations, and alterations in neuromuscular function associated with extended sleep that impact muscle strength.^4,52^ For instance, extended sleep could interfere with the natural rhythm of anabolic and catabolic hormone secretion, which is crucial for muscle recovery and strength.^53^ Sleep also affects other hormones like cortisol is involved in muscle protein degradation.^54,55^ Previous studies have associated longer sleep duration to various adverse health-related outcomes, and we found its association with lower muscle strength. The explanation for these associations remains to be elucidated.^56,57^

Our analysis boasts strengths in two key areas. Firstly, a diverse cohort of both young and older adults ensures broad applicability of findings. Secondly, the study uniquely isolates lifestyle factors’ independent impact on muscle mass and strength, offering a comprehensive view. Despite strengths, limitations include the cross-sectional design limiting causal inferences and potential bias from self-reported data, necessitating objective measures like accelerometers. Another limitation, unexplored variables such as genetics could enhance understanding. The study uncovers the intricate relationship between lifestyle and aging muscle health, noting the nuanced interplay of muscle mass and strength. Men’s muscle health is linked to dietary fiber intake, water consumption, and physical activity, while women show associations with dietary fiber intake and physical activity. For women, only dietary fiber intake and physical activity exhibit this association, affirming the efficacy of physical activity and suggesting the gut-muscle axis’s underestimated role in aging. This research may pave new ways for promoting healthier aging and sustaining muscle health.

## CONCLUSION

Our study emphasizes lifestyle factors influencing muscle strength over muscle mass. Physical activity and fiber intake, however, affect both outcomes in both genders across ages up to 70 years, potentially involving the gut-muscle axis. Our findings in stratified analyses underscore the complex nature of muscle health, requiring longitudinal research to provide personalized lifestyle recommendations to specific populations.

## Data Availability

All data produced in the present study are available upon reasonable request to the authors

## AUTHOR CONTRIBUTIONS

JMM and MB conceptualized the study, JMM drafted outline and the first version of the manuscript. MRG, MB, CSP, SV, and PS provided methodological support. All authors had full access to all the data in the study, discussed the results, and had final responsibility for the decision to submit for publication. JMM and MB accessed and verified the data.

## ACKNOWLEDGMENTS

This research has been conducted using the UK Biobank Resource under Application Number 78275.

## REFERENCES

1. Kanasi, E., Ayilavarapu, S. & Jones, J. The aging population: demographics and the biology of aging. Periodontol 2000 72, 13–18 (2016).

2. Janssen, I., Heymsfield, S. B., Wang, Z. M. & Ross, R. Skeletal muscle mass and distribution in 468 men and women aged 18-88 yr. J Appl Physiol (1985) 89, 81–88 (2000).

3. Krzysztofik, M., Wilk, M., Wojdała, G. & Gołaś, A. Maximizing Muscle Hypertrophy: A Systematic Review of Advanced Resistance Training Techniques and Methods. Int J Environ Res Public Health 16, (2019).

4. Genario, R. et al. Sleep quality is a predictor of muscle mass, strength, quality of life, anxiety and depression in older adults with obesity. Scientific Reports 2023 13:1 13, 1–9 (2023).

5. Carbone, J. W. & Pasiakos, S. M. Dietary Protein and Muscle Mass: Translating Science to Application and Health Benefit. Nutrients 11, (2019).

6. Cui, Y. et al. The longitudinal association between alcohol consumption and muscle strength: A population-based prospective study. J Musculoskelet Neuronal Interact 19, 294 (2019).

7. Serra-Prat, M., Lorenzo, I., Palomera, E., Ramírez, S. & Yébenes, J. C. Total Body Water and Intracellular Water Relationships with Muscle Strength, Frailty and Functional Performance in an Elderly Population. J Nutr Health Aging 23, 96–101 (2019).

8. Frampton, J., Murphy, K. G., Frost, G. & Chambers, E. S. Higher dietary fibre intake is associated with increased skeletal muscle mass and strength in adults aged 40 years and older. J Cachexia Sarcopenia Muscle 12, 2134 (2021).

9. Morwani-Mangnani, J. et al. Gut microbiome changes due to sleep disruption in older and younger individuals: a case for sarcopenia? Sleep 45, (2022).

10. Auyeung, T. W. et al. Sleep Duration and Disturbances Were Associated With Testosterone Level, Muscle Mass, and Muscle Strength—A Cross-Sectional Study in 1274 Older Men. J Am Med Dir Assoc 16, 630.e1–630.e6 (2015).

11. Sudlow, C. et al. UK Biobank: An Open Access Resource for Identifying the Causes of a Wide Range of Complex Diseases of Middle and Old Age. PLoS Med 12, e1001779 (2015).

12. Townsend, P., Phillimore, P. & Beattie, A. Health and Deprivation: Inequality and the North. Preprint at (1988).

13. Noble, M., Wright, G., Smith, G. & Dibben, C. Measuring multiple deprivation at the small-area Level. Environ Plan A 38, 169–185 (2006).

14. Lloyd, C. D., Norman, P. D. & McLennan, D. Deprivation in England, 1971–2020. Appl Spat Anal Policy 16, 461–484 (2023).

15. Willett, W. C., Howe, G. R. & Kushi, L. H. Adjustment for total energy intake in epidemiologic studies. Am J Clin Nutr 65, (1997).

16. Chen, L. K. et al. Asian Working Group for Sarcopenia: 2019 Consensus Update on Sarcopenia Diagnosis and Treatment. J Am Med Dir Assoc 21, 300–307.e2 (2020).

17. Landi, F. et al. Anorexia of Aging: Risk Factors, Consequences, and Potential Treatments. Nutrients 8, (2016).

18. Paterson, D. H. et al. Ageing and physical activity: evidence to develop exercise recommendations for older adultsThis article is part of a supplement entitled Advancing physical activity measurement and guidelines in Canada: a scientific review and evidence-based foundation for the future of Canadian physical activity guidelines co-published by Applied Physiology, Nutrition, and Metabolism and the Canadian Journal of Public Health. It may be cited as Appl. Physiol. Nutr. Metab. 32(Suppl. 2E) or as Can. J. Public Heal. 10.1139/H07-111 32, (2007).

19. Nuzzo, J. L. Narrative Review of Sex Differences in Muscle Strength, Endurance, Activation, Size, Fiber Type, and Strength Training Participation Rates, Preferences, Motivations, Injuries, and Neuromuscular Adaptations. J Strength Cond Res 37, 494– 536 (2023).

20. Yu, F., Hedström, M., Cristea, A., Dalén, N. & Larsson, L. Effects of ageing and gender on contractile properties in human skeletal muscle and single fibres. Acta Physiologica 190, 229–241 (2007).

21. Barone, B. et al. The Role of Testosterone in the Elderly: What Do We Know? International Journal of Molecular Sciences 2022, Vol. 23, Page 3535 23, 3535 (2022).

22. Esbjörnsson, M. E., Dahlström, M. S., Gierup, J. W. & Jansson, E. C. Muscle fiber size in healthy children and adults in relation to sex and fiber types. Muscle Nerve 63, 586– 592 (2021).

23. Schoenfeld, B. J., Grgic, J., Ogborn, D. & Krieger, J. W. Strength and Hypertrophy Adaptations Between Low- vs. High-Load Resistance Training: A Systematic Review and Meta-analysis. J Strength Cond Res 31, 3508–3523 (2017).

24. Del Vecchio, A. et al. The increase in muscle force after 4 weeks of strength training is mediated by adaptations in motor unit recruitment and rate coding. J Physiol 597, 1873–1887 (2019).

25. Rom, O., Kaisari, S., Aizenbud, D. & Reznick, A. Z. Lifestyle and Sarcopenia— Etiology, Prevention, and Treatment. Rambam Maimonides Med J 3, e0024 (2012).

26. Vaz, S. et al. Prescribing tailored home exercise program to older adults in the community using a tailored self-modeled video: A pre-post study. Front Public Health 10, (2022).

27. Li, G., Li, X. & Chen, L. Personally tailored exercises for improving physical outcomes for older adults in the community: A systematic review. Arch Gerontol Geriatr 101, (2022).

28. Hwang, J. & Park, S. Gender-Specific Risk Factors and Prevalence for Sarcopenia among Community-Dwelling Young-Old Adults. Int J Environ Res Public Health 19, (2022).

29. Du, Y. et al. Sex differences in the prevalence and adverse outcomes of sarcopenia and sarcopenic obesity in community dwelling elderly in East China using the AWGS criteria. BMC Endocr Disord 19, 1–11 (2019).

30. Tay, L. et al. Sex-specific differences in risk factors for sarcopenia amongst community-dwelling older adults. Age (Omaha) 37, 1–12 (2015).

31. Prokopidis, K., Chambers, E., Ni Lochlainn, M. & Witard, O. C. Mechanisms Linking the Gut-Muscle Axis With Muscle Protein Metabolism and Anabolic Resistance: Implications for Older Adults at Risk of Sarcopenia. Front Physiol 12, 770455 (2021).

32. Ticinesi, A. et al. Aging gut microbiota at the cross-road between nutrition, physical frailty, and sarcopenia: Is there a gut–muscle axis? Nutrients 9, 1–20 (2017).

33. Ticinesi, A. et al. Exercise and immune system as modulators of intestinal microbiome: Implications for the gut-muscle axis hypothesis. Exerc Immunol Rev 25, 84–95 (2019).

34. Taniguchi, H. et al. Effects of short-term endurance exercise on gut microbiota in elderly men. Physiol Rep 6, 13935 (2018).

35. Frampton, J., Murphy, K. G., Frost, G. & Chambers, E. S. Short-chain fatty acids as potential regulators of skeletal muscle metabolism and function. Nat Metab 2, 840–848 (2020).

36. Chen, F. et al. Association of the gut microbiota and fecal short-chain fatty acids with skeletal muscle mass and strength in children. The FASEB Journal 36, e22109 (2022).

37. Frampton, J., Murphy, K. G., Frost, G. & Chambers, E. S. Higher dietary fibre intake is associated with increased skeletal muscle mass and strength in adults aged 40 years and older. J Cachexia Sarcopenia Muscle 12, 2134 (2021).

38. Montiel-Rojas, D. et al. Dietary Fibre May Mitigate Sarcopenia Risk: Findings from the NU-AGE Cohort of Older European Adults. Nutrients 12, (2020).

39. Frampton, J., Murphy, K. G., Frost, G. & Chambers, E. S. Higher dietary fibre intake is associated with increased skeletal muscle mass and strength in adults aged 40 years and older. J Cachexia Sarcopenia Muscle 12, 2134 (2021).

40. Grandjean, A. NUTRITIONAL REQUIREMENTS TO INCREASE LEAN MASS. Clin Sports Med 18, 623–632 (1999).

41. Kerksick, C. M. et al. International society of sports nutrition position stand: nutrient timing. J Int Soc Sports Nutr 14, (2017).

42. Højfeldt, G. et al. Daily Protein and Energy Intake Are Not Associated with Muscle Mass and Physical Function in Healthy Older Individuals—A Cross-Sectional Study. Nutrients 12, 1–16 (2020).

43. Verreijen, A. M. et al. Dietary protein intake is not associated with 5-y change in mid-thigh muscle cross-sectional area by computed tomography in older adults: the Health, Aging, and Body Composition (Health ABC) Study. Am J Clin Nutr 109, 535–543 (2019).

44. Evans, E. M. et al. Effects of protein intake and gender on body composition changes: a randomized clinical weight loss trial. Nutr Metab (Lond) 9, 55 (2012).

45. Lorenzo, I., Serra-Prat, M. & Carlos Yébenes, J. The Role of Water Homeostasis in Muscle Function and Frailty: A Review. Nutrients 11, (2019).

46. Fernando, S., Sivagnanam, F. & Rathish, D. A compulsive act of excess water intake leading to hyponatraemia and rhabdomyolysis: a case report. Int J Emerg Med 12, (2019).

47. Kim, H., Beom, S. H., Kim, T. H. & Kim, B. J. Association of Water Intake with Hand Grip Strength in Community-Dwelling Older Adults. Nutrients 13, (2021).

48. Collins, M. A. et al. Alcohol in Moderation, Cardioprotection and Neuroprotection: Epidemiological Considerations and Mechanistic Studies. Alcohol Clin Exp Res 33, 206 (2009).

49. Kojima, G., Iliffe, S., Liljas, A. & Walters, K. Non-linear association between alcohol and incident frailty among community-dwelling older people: A dose-response meta-analysis. Biosci Trends 11, 600–602 (2017).

50. White, A. M. Gender Differences in the Epidemiology of Alcohol Use and Related Harms in the United States. Alcohol Res 40, 1–13 (2020).

51. Skinner, J., Shepstone, L., Hickson, M. & Welch, A. A. Alcohol Consumption and Measures of Sarcopenic Muscle Risk: Cross-Sectional and Prospective Associations Within the UK Biobank Study. Calcif Tissue Int 113, 143–156 (2023).

52. Kim, H. J. et al. Sex differences in deterioration of sleep properties associated with aging: a 12-year longitudinal cohort study. J Clin Sleep Med 17, 964 (2021).

53. Dáttilo, M. et al. Effects of Sleep Deprivation on Acute Skeletal Muscle Recovery after Exercise. Med Sci Sports Exerc 52, 507–514 (2020).

54. Lamon, S. et al. The effect of acute sleep deprivation on skeletal muscle protein synthesis and the hormonal environment. Physiol Rep 9, (2021).

55. Auyeung, T. W. et al. Sleep Duration and Disturbances Were Associated With Testosterone Level, Muscle Mass, and Muscle Strength--A Cross-Sectional Study in 1274 Older Men. J Am Med Dir Assoc 16, 630.e1–630.e6 (2015).

56. Wang, D., Ruan, W., Peng, Y. & Li, W. Sleep duration and the risk of osteoporosis among middle-aged and elderly adults: a dose-response meta-analysis. Osteoporosis International 29, 1689–1695 (2018).

57. Ohara, T. et al. Association Between Daily Sleep Duration and Risk of Dementia and Mortality in a Japanese Community. J Am Geriatr Soc 66, 1911–1918 (2018).

