## Supplemental Figure 1 for "Physical activity and fiber intake beneficial for muscle mass and strength preservation during aging: A Comprehensive Cross-Sectional Study in the UK Biobank cohort"

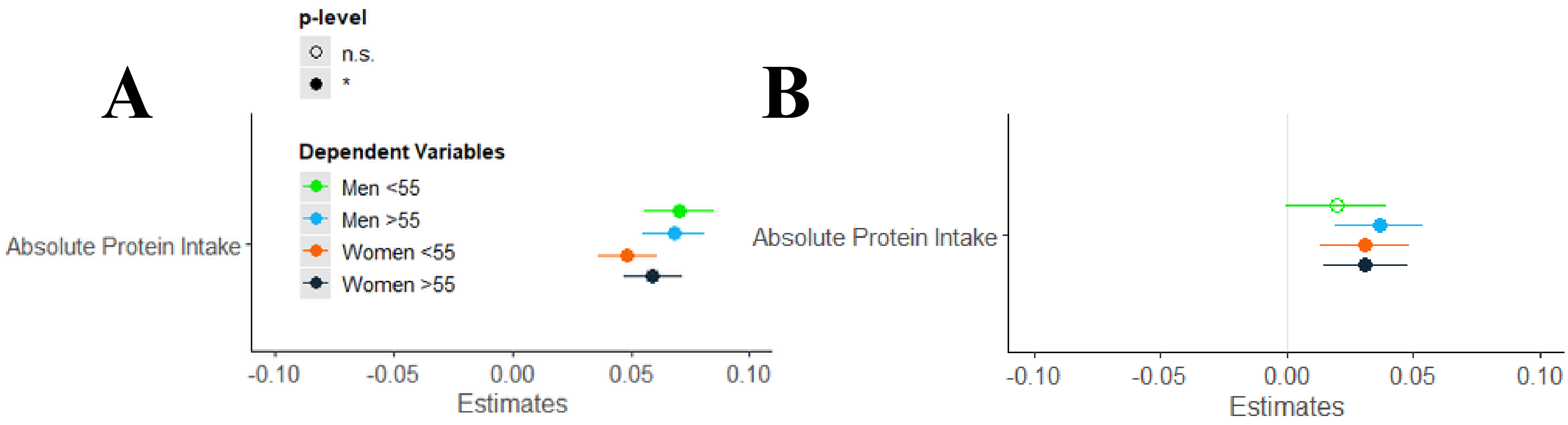


**FIGURE S1. Absolute protein intake (not-energy adjusted) in the multivariate model presented as standardized beta-coefficients associated with (A) muscle mass and (B) handgrip strength for men and women younger and older than 55 years**. Adjusted for traditional covariates, socio-demographic covariates, medication use, and smoking status. The p-value at which to accept significance (filled in circle) was adjusted to account for multiple testing ($p < 0.0036$).
